# Risk factors and outcomes associated with heart failure with preserved and reduced ejection fraction in persons with chronic kidney disease

**DOI:** 10.1101/2023.08.24.23294596

**Authors:** Nisha Bansal, Leila Zelnick, Rebecca Scherzer, Michelle Estrella, Michael Shlipak

**Author notes:** Corresponding author: Nisha Bansal MD MAS Division of Nephrology, University of Washington 908 Jefferson St, 3^rd^ floor Seattle, WA 98104 Facsimile.

## Abstract

**Background:** Heart failure (HF) is associated with poor outcomes in persons with chronic kidney disease (CKD), yet there are limited data on whether outcomes differ by HF subtype. This study aimed to examine associations of incident preserved (HFpEF) versus reduced (HFrEF) ejection fraction (EF) with risk of progression to end-stage-kidney-disease (ESKD) and mortality.

**Methods:** We studied individuals with mild to severe CKD enrolled in the Chronic Renal Insufficiency Cohort (CRIC) study who were free of HF at cohort entry. Incident HF hospitalizations were adjudicated during study follow-up and classified into HFpEF (EF≥50%) or HFrEF (EF<50%) based on echocardiograms performed during the hospitalization or at a research study visit within one year of the hospitalization. ESKD was defined as need for chronic dialysis or kidney transplant during follow-up. Cox proportional hazards were used to evaluate the association of time-updated HF subtype with risk of ESKD and mortality, adjusting for demographics, comorbidities and medication use.

**Results:** Among the 3,557 study participants without HF at cohort entry, mean age was 57 years and mean eGFR 45 ml/min/1.73 m^2^ at baseline. Incidence rates for HFpEF and HFrEF were 0.9 (95% CI 0.8, 1.0) and 0.7 (95% CI 0.6, 0.8) per 100 person-years, respectively. Adjusted associations of incident HF with progression to ESKD were similar for HFpEF (HR 1.59, 95% CI: 1.24, 2.02) and HFrEF (HR 1.26, 95% CI: 0.93, 1.70) (test for difference p-value=0.35). The adjusted associations of HFpEF and HFrEF HF with mortality were stronger for HFrEF (HR 1.68, 95% CI: 1.34, 2.11) compared with HFpEF (HR 1.24, 95% CI: 1.00, 1.54) (test for difference p-value = 0.02).

**Conclusions:** In a large U.S. CKD population, the rates of HFpEF hospitalizations were greater than that of HFrEF. Both types of HF had similar associations with risk of ESKD; however, there was a stronger association of HFrEF with mortality. Prevention and treatment of both HFpEF and HFrEF should be central priorities to improve clinical outcomes in patients with CKD.

## INTRODUCTION

Heart failure (HF) is one of the leading causes of morbidity and mortality among patients with chronic kidney disease (CKD), ^1-6^ with unique risk factors and pathophysiology compared with the general population. The physiological relationships between CKD and HF are multifactorial and intertwined. For example, CKD contributes to HF by increased salt retention and volume expansion, upregulation of neurohormonal pathways, and proinflammatory mechanisms among other disturbances.^7^ Patients with CKD have an estimated threefold risk of incident HF compared to those without CKD.^2^ Studies from our group and others have demonstrated that persons with CKD have a substantially higher risk of mortality and CKD progression after developing HF, compared to those who have not developed HF.^8^

Distinctions between HF with preserved ejection fraction (HFpEF) and HF with reduced ejection fraction (HFrEF) have been recognized, including differing pathophysiology and consequent treatment approaches.^9,10^ A prior study reported that among patients with CKD, the incidence and prevalence of HFpEF are higher than that of HFrEF.^8^ However, the relative contributions of various risk factors to each of these HF subtypes have not been well characterized, and these etiological differences may have important implications for prognosis and treatment. Therefore, the aim of this study was to compare risk factors for HFpEF versus HFrEF in a large, national CKD population, and to examine associations of HFpEF and HFrEF with risk of mortality and progression to end-stage-kidney-disease (ESKD), two important outcomes for patients with CKD.

## METHODS

### Study population

We studied individuals with mild to severe CKD enrolled in the Chronic Renal Insufficiency Cohort (CRIC) study, with data from the National Institute of Diabetes and Digestive and Kidney Diseases (NIDDK) public repository. The CRIC study recruited 3,939 participants with mild to severe CKD, defined as an eGFR 20-70 ml/min/1.73m^2^, between June 2003 and August 2008 at seven clinical centers across the U.S. (Ann Arbor/Detroit, MI; Baltimore, MD; Chicago, IL; Cleveland, OH; New Orleans, LA; Philadelphia, PA; and Oakland, CA).^11,12^ All study participants provided written informed consent, and the study protocol was approved by institutional review boards at each of the participating sites. Detailed inclusion and exclusion criteria have been previously described.^11^ Participants on maintenance dialysis or with a kidney transplant were not included at cohort entry. CRIC also excluded participants with advanced HF, defined as New York Heart Association Class III or IV, on cohort entry. For the present study, we excluded 382 participants who had a history of HF at baseline, resulting in a final analytical cohort of 3,557 participants free of HF at cohort entry.

All participants enrolled in the study had annual in-person study visits where detailed interviews were conducted and brief physical examination, laboratory measures and cardiovascular testing were performed. In addition to the annual study visits, all CRIC participants were contacted every 6 months to obtain updated information on medication use and interim updates to medical history/hospitalizations.

### Ascertainment of heart failure hospitalizations and heart failure subtype

Incident HF events were based on hospitalizations for HF which were adjudicated from study entry through December 2019. Hospitalizations for HF were identified by asking study participants semi-annually if they were hospitalized and by electronic health record queries of selected hospitals or health care systems for qualifying encounters. The first 30 discharge codes were identified for all hospitalizations, and codes relevant to HF resulted in retrieval of medical records by study personnel for centralized adjudicated review. At least two study physicians reviewed all possible HF events and deaths using medical records and adjudicated for clinical HF based on clinical symptoms, radiographic evidence of pulmonary congestion, physical examination of the heart and lungs and, when available, central venous hemodynamic monitoring data, and echocardiographic imaging. HF was confirmed when both reviewers agreed upon a “probable” or “definite” occurrence of HF.

We stratified HF by preserved and reduced ejection fraction HF (HFpEF and HFrEF). HFpEF was defined as ejection fraction ≥50%, and HFrEF was defined as ejection fraction <50%. Given the time-updated approach, participants were able to change HF subtype categorization during follow-up. Ejection fraction was ascertained from echocardiograms performed during the index hospitalization for clinical purposes. If an echocardiogram was not performed during the index hospitalization, we utilized the ejection fraction quantified from an ambulatory CRIC research echocardiogram up to 1 year before or after the index HF hospitalization. Research echocardiograms in CRIC were performed at multiple time-points including years 1, 4, 7 as well as when the participant progressed to eGFR<20 ml/min/1.73m^2^. Our previous work has shown that ejection fraction in CRIC is generally stable over time.^13,14^ Among a total of 682 incident HF hospitalizations, 521 (76%) had ejection fraction available through either a clinical echocardiogram during the index hospitalization or a CRIC research echocardiogram. Otherwise, they were classified as “unspecified HF.”

### Definition of progression to end-stage-kidney-disease (ESKD)

ESKD was defined as receipt of chronic dialysis or a kidney transplant and was identified through participant self-report, medical records review and data from the USRDS.

### All-cause mortality

Death status was ascertained from study entry through end of follow-up. Deaths were identified from report from next of kin, retrieval of death certificates or obituaries, review of hospital or outpatient records, and linkage to Social Security Death vital status and state death certificate files, if available.

### Covariates

All covariates were obtained from the baseline study visit and time-updated if a participant changed HF status. At the baseline and each annual study visit, participants provided information on their sociodemographic characteristics, medical history, medication usage, and lifestyle behaviors. Race and ethnicity were categorized as non-Hispanic white, non-Hispanic Black, Hispanic, and other. History of cardiovascular disease was determined by self-report and included history of HF, myocardial infarction, coronary revascularization or stroke. Anthropometric measurements and blood pressure (BP) were assessed using standard protocols.^15^ Metabolic equivalent of task (MET) minutes, a measure of physical activity, were calculated based on report by study participants.

Estimated glomerular filtration rate (eGFR) and urine albumin-to-creatinine ratio (urine ACR) were measured via standardized methods at annual research study visits. Serum creatinine was measured using an enzymatic method on an Ortho Vitros 950 at the CRIC central laboratory and standardized to isotope dilution mass spectrometry-traceable values.^16,17^ Laboratory tests including: electrolytes, lipids, inflammatory markers (IL-6 and C-reactive protein), galectin-3, growth differentiation factor (GDF)-15, soluble ST2 (sST2), FGF-23 and hemoglobin A1C were measured at the baseline visit. Estimated GFR was calculated using serum creatinine and serum cystatin C with the 2021 combined CKD-EPI equation^18^ and categorized as ≥45, 30-44 and <30 ml/min/1.73 m^2^.^19^ Urine ACR was quantified from 24-hour urine samples and categorized as <30, 30-299 or>300 mg/g.^19^

Body mass index (BMI) was calculated as weight in kg divided by height in meters squared. Diabetes mellitus was defined as a fasting glucose >126 mg/dL, a non-fasting glucose >200 mg/dL, or use of insulin or other antidiabetic medication. Cardiovascular medications, including diuretics, angiotensin converting enzyme (ACE) inhibitors/angiotensin receptor blockers (ARBs), and β-blockers, were ascertained by detailed review with participants at baseline.

### Statistical analyses

We first described characteristics of study participants in our cohort. Participants were censored at death, loss to follow-up, study withdrawal or end of study follow-up (through December 2019), whichever came first. Over the follow-up period, we calculated the unadjusted rates (per 100 person-years) of HF hospitalizations overall and by HF subtype (HFpEF, HFrEF or unspecified) overall and by eGFR categories (≥45, 30-44 and <30 ml/min/1.73m^2^); 95% confidence intervals (CI) were obtained using non-parametric bootstrap methods. We utilized multivariable Cox regression with Wei, Lin, and Weissfeld standard errors^20^ to examine the association of participant characteristics with risk of the first HF subtype during follow-up, and to test differences in risk factor associations between HF subtypes.

Kaplan-Meier curves were generated to model the ESKD-free survival and mortality after diagnosis of either HFpEF or HFrEF overall and by eGFR category. Cox regression with a time-updated exposure of HF subtype were performed to test the association of HF subtype with the outcomes of interest, progression to ESKD and mortality. With the time-updated approach, all HF hospitalizations were included as exposures in the model, and participants may have had multiple HF events of different HF subtypes. In this approach, participants could contribute time at risk for multiple HF hospitalizations, and subsequent follow-up time began immediately following HF hospitalization for the outcomes of ESKD or death. Covariates were obtained from the closest study visit prior to the hospitalization. We adjusted for age, sex, race, ethnicity, diabetes, history of cardiovascular disease, atrial fibrillation, current smoking, systolic blood pressure, BMI and for use of cardiovascular medications (diuretics, ACEi/ARBs and β-blockers).

In a secondary analysis, we used Cox regression models with a similar approach as the primary analyses to examine the association of eGFR categories (<30, 30-44 and >45 ml/min/1.73m^2^) with risk of ESKD and mortality among participants who developed incident HFpEF or HFrEF.

All analyses were performed using R 4.2.3 (R Foundation for Computing, Vienna, Austria).

## RESULTS

### Characteristics of study population

Among the 3,557 study participants without HF at cohort entry, mean age was 57 years, 43% identified as white race, prevalence of diabetes was 46%, history of myocardial infarction was 18%, mean systolic blood pressure was 129 mm Hg, and mean eGFR 45 ml/min/1.73 m^2^ (**Supplemental Table 1**).

### Rates and risk factors for HFpEF and HFrEF

A total of 682 participants had an incident HF hospitalization during follow-up, of whom 443 had only one HF classification (HFpEF, HFrEF or unspecified) whereas 239 participants had more than one HF subtype over the follow-up period. Median time to first HFpEF hospitalization was 4.7 [IQR 1.7-8.4] years and for HFrEF 5.0 [IQR 2.6-9.2] years from baseline. The incidence rate of HF overall was 2.1 (95% CI 1.9, 2.2) per 100 person-years. Incidence rates for HFpEF and HFrEF were 0.9 (95% CI 0.8, 1.0) and 0.7 (95% CI 0.6, 0.8) per 100 person-years, respectively. Rates of HFpEF were higher than HFrEF in all eGFR categories (**Figure 1**).

**Figure 1.**
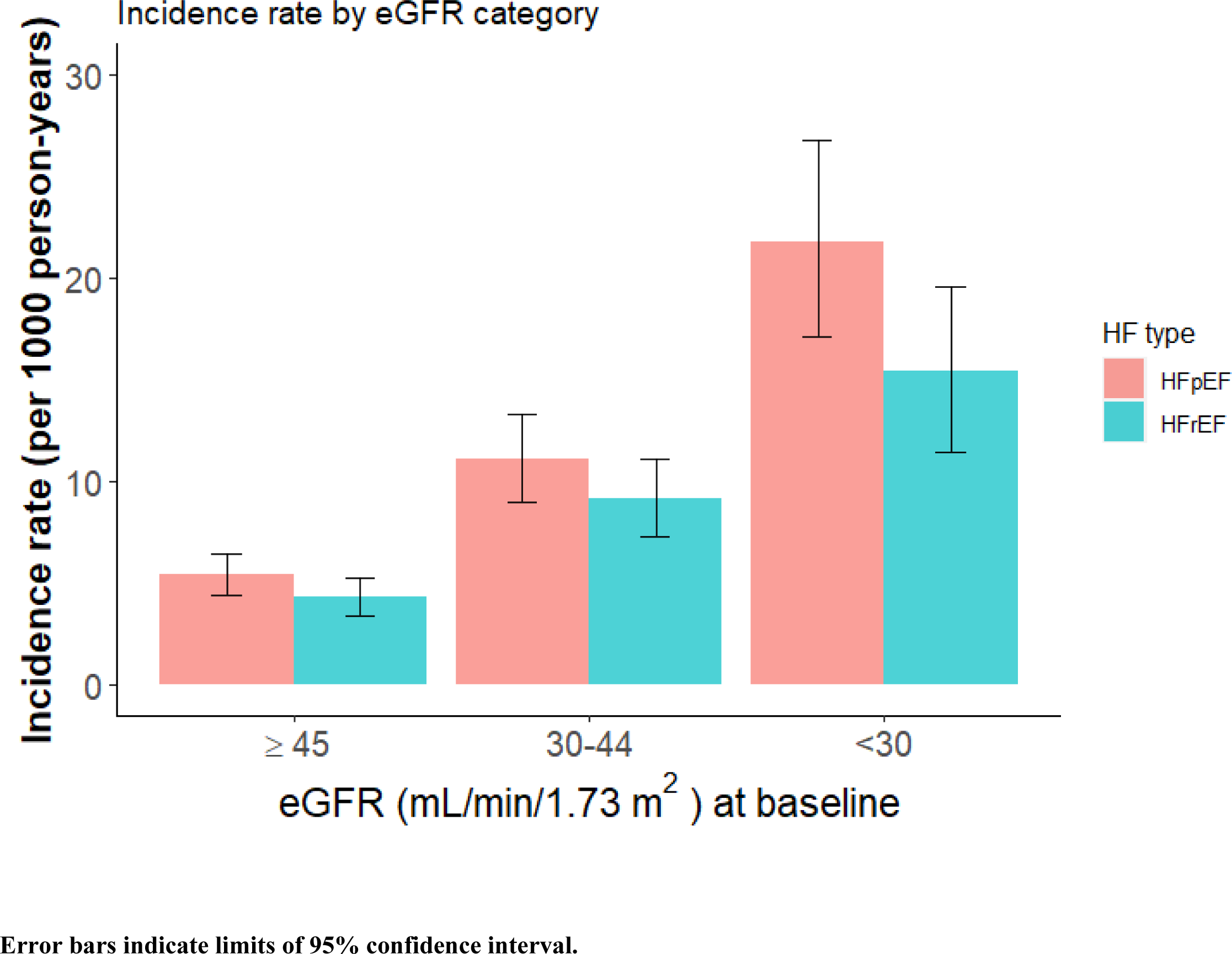
Incidence rates of heart failure with preserved (HFpEF) and reduced (HFrEF) ejection fraction by estimated glomerular filtration rate (eGFR) category.

In comparing risk factors for HF subtypes, older age, higher urine ACR, and history of atrial fibrillation and myocardial infarction were associated with risk of both HFpEF and HFrEF. Certain risk factors did differ for HFpEF versus HFrEF (**Table 1**). Higher systolic blood pressure, lower diastolic blood pressure, higher hemoglobin A1C, higher FGF-23, higher IL-6 and higher GDF-15 were more strongly associated with HFpEF. Male sex, higher heart rate, history of COPD, smoking, lower eGFR, higher BMI, and lower waist circumference were more strongly associated with HFrEF. Statistically significant interactions by HF subtype were observed for male sex, blood pressure, heart rate and waist circumference.

**Table 1.** Multivariable association of baseline patient characteristics and risk factors with the first heart failure event among participants with chronic kidney disease.

|  | Heart failure with<br>preserved ejection<br>fraction | Heart failure with<br>reduced ejection<br>fraction |  |
| --- | --- | --- | --- |
| Variable | HR (95% CI) | HR (95% CI) | p for<br>interaction |
| Age (per 10 year increment) | **1.24 (1.05, 1.46)** | **1.50 (1.25, 1.79)** | 0.13 |
| Male | 1.00 (0.73, 1.37) | **1.78 (1.21, 2.61)** | **0.02** |
| Race/ethnicity |  |  |  |
| White | 1.0 (Ref.) | 1.0 (Ref.) |  |
| Black | 1.27 (0.92, 1.75) | 0.87 (0.60, 1.26) | 0.13 |
| Hispanic | 0.71 (0.45, 1.10) | 0.73 (0.46, 1.18) | 0.92 |
| Other | 1.20 (0.66, 2.20) | 0.72 (0.30, 1.72) | 0.34 |
| Atrial fibrillation | **1.41 (1.04, 1.91)** | **1.82 (1.32, 2.51)** | 0.25 |
| Diabetes | 1.06 (0.73, 1.54) | 1.11 (0.72, 1.71) | 0.89 |
| Myocardial infarction | **1.61 (1.22, 2.12)** | **2.22 (1.64, 3.01)** | 0.12 |
| Peripheral vascular disease | 0.96 (0.64, 1.46) | 1.19 (0.76, 1.86) | 0.51 |
| Stroke | 1.31 (0.92, 1.85) | 0.94 (0.60, 1.46) | 0.25 |
| Current smoking | 1.25 (0.87, 1.79) | **1.77 (1.22, 2.57)** | 0.19 |
| eGFR (per 15 mL/min/1.73m<sup>2</sup><br>decrement) | 1.05 (0.87, 1.27) | **1.27 (1.04, 1.55)** | 0.16 |
| Urine albumin-to-creatinine ratio<br>(24hour) (per SD increment in log) | **1.34 (1.12, 1.61)** | **1.45 (1.19, 1.77)** | 0.58 |
| Body mass index (per 5 kg/m<sup>2</sup><br>increment) | 1.03 (0.88, 1.20) | **1.21 (1.04, 1.41)** | 0.13 |
| Systolic blood pressure (per 10<br>mmHg increment) | **1.14 (1.06, 1.22)** | 1.00 (0.92, 1.09) | **0.02** |
| Diastolic blood pressure (per 10<br>mmHg increment) | **0.81 (0.71, 0.93)** | 1.10 (0.93, 1.30) | **0.005** |
| Chronic obstructive pulmonary<br>disease | 1.10 (0.57, 2.15) | **2.16 (1.22, 3.83)** | 0.13 |
| Hemoglobin (per SD decrement) | 1.08 (0.90, 1.28) | 1.21 (0.98, 1.49) | 0.40 |
| Glucose (per SD increment) | 0.98 (0.87, 1.11) | 0.95 (0.82, 1.10) | 0.76 |
| Serum albumin (per SD<br>decrement) | 0.92 (0.78, 1.09) | 0.98 (0.81, 1.17) | 0.65 |
| Serum sodium (per SD increment) | 0.91 (0.80, 1.05) | 0.96 (0.82, 1.12) | 0.63 |
| Calcium (per SD decrement) | **1.19 (1.01, 1.40)** | 1.14 (0.94, 1.38) | 0.78 |
| Phosphate (per SD increment) | 1.08 (0.95, 1.22) | 0.92 (0.78, 1.07) | 0.11 |
| Parathyroid hormone (per SD | 0.97 (0.83, 1.12) | 1.04 (0.86, 1.24) | 0.56 |
| Variable | HR (95% CI) | HR (95% CI) | p for<br>interaction |
| increment in log) |  |  |  |
| Heart rate (per SD increment) | 0.96 (0.84, 1.10) | **1.25 (1.06, 1.46)** | **0.02** |
| Waist circumference (per SD<br>increment) | 1.26 (0.99, 1.59) | **0.75 (0.57, 0.98)** | **0.004** |
| Alcohol use | 1.21 (0.93, 1.57) | 0.97 (0.72, 1.31) | 0.28 |
| Triglycerides (per SD increment) | 1.03 (0.90, 1.19) | 1.00 (0.88, 1.14) | 0.73 |
| Total cholesterol (per SD<br>increment) | 1.01 (0.88, 1.16) | 1.08 (0.91, 1.29) | 0.55 |
| HbA1c (per SD increment) | **1.30 (1.12, 1.51)** | 1.19 (0.97, 1.46) | 0.48 |
| FGF-23 (per SD increment in log) | **1.19 (1.01, 1.39)** | 1.11 (0.94, 1.31) | 0.57 |
| High sensitivity C-reactive protein<br>(per SD increment in log) | 1.00 (0.87, 1.16) | 1.05 (0.90, 1.23) | 0.65 |
| IL-6 (per SD increment in log) | **1.16 (1.01, 1.34)** | 1.13 (0.98, 1.31) | 0.78 |
| Total METs (per SD decrement) | 0.97 (0.84, 1.12) | 1.10 (0.92, 1.32) | 0.28 |
| Galectin-3 (per SD increment in<br>log) | 1.07 (0.92, 1.25) | 1.05 (0.89, 1.24) | 0.87 |
| GDF-15 (per SD increment in log) | **1.37 (1.13, 1.67)** | 1.01 (0.80, 1.29) | 0.06 |
| sST2 (per SD increment in log) | 1.12 (0.98, 1.29) | 1.07 (0.92, 1.24) | 0.63 |
| 24 hour urine sodium (per SD<br>decrement) | 1.06 (0.91, 1.22) | 1.03 (0.88, 1.21) | 0.84 |
| 24 hour urine potassium (per SD<br>decrement) | 0.96 (0.82, 1.12) | 0.84 (0.69, 1.01) | 0.30 |

### Association of HFpEF and HFrEF with progression to ESKD

ESKD-free survival probabilities decreased rapidly over time for both HFpEF and HFrEF (**Figures 2a and 2b).** The median time to ESKD among patients with HFpEF and HFrEF were 1.1 (IQR 0.1-2.4) and 1.1 (0.2-3.1) years, respectively. In unadjusted models, incident HFpEF and HFrEF were associated with 5-fold greater risk of progression to ESKD compared with no HF (**Table 2**). When adjusted for demographic characteristics, comorbidity, measures of kidney function and medication use, the associations of incident HFpEF (HR 1.59 [95% CI: 1.24, 2.02]) with progression to ESKD remained statistically significant; while the association of incident HFrEF with ESKD was no longer statistically significant (HR 1.26, 95% CI, 0.93, 1.70). Associations of HFpEF and HFrEF with ESKD risk were not statistically different (p-value=0.35).

**Figure 2.**
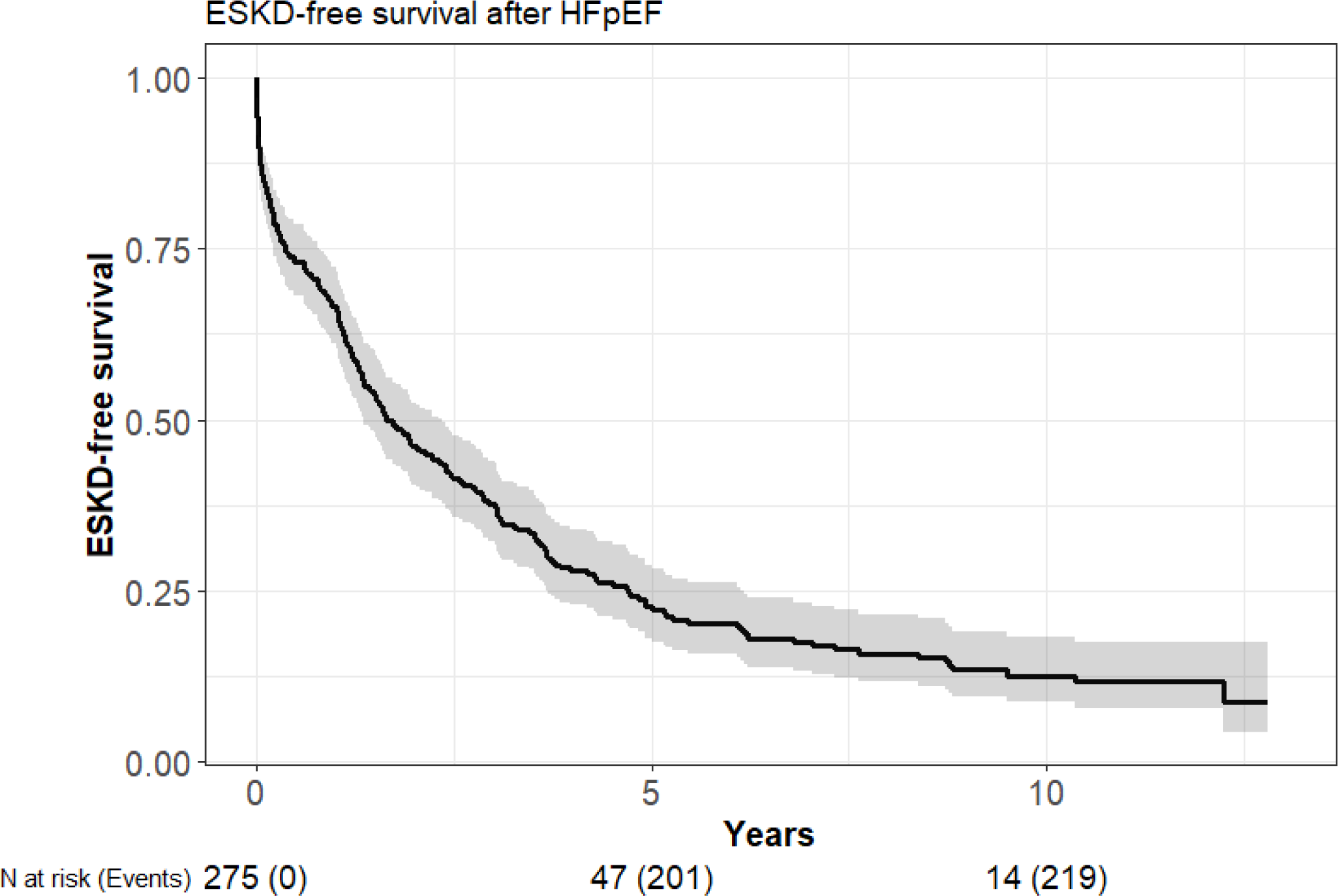
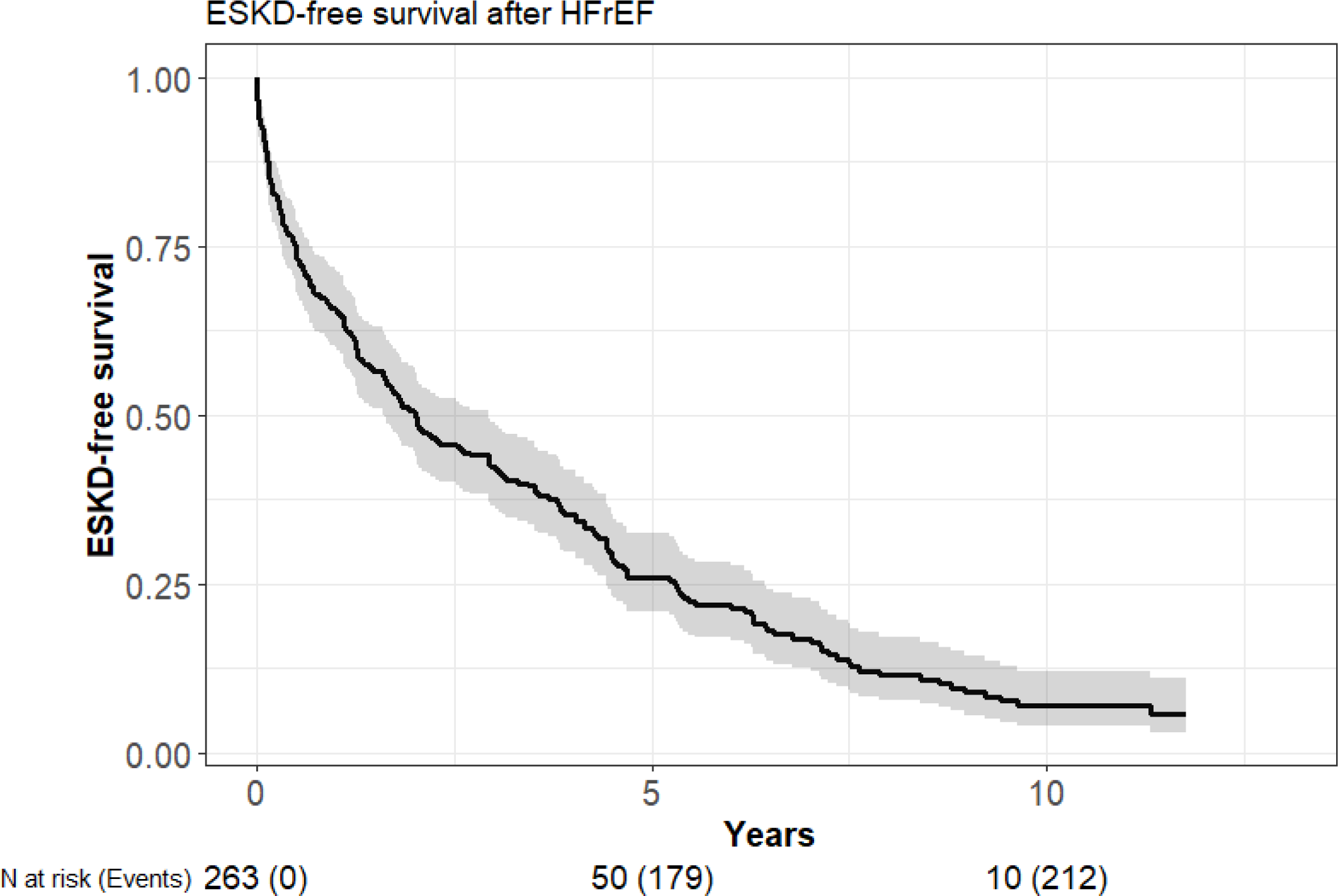
End-stage kidney disease (ESKD) free survival after incident heart failure with (a) preserved ejection fraction (HFpEF) and (b) reduced ejection fraction (HFrEF)

**Table 2.**
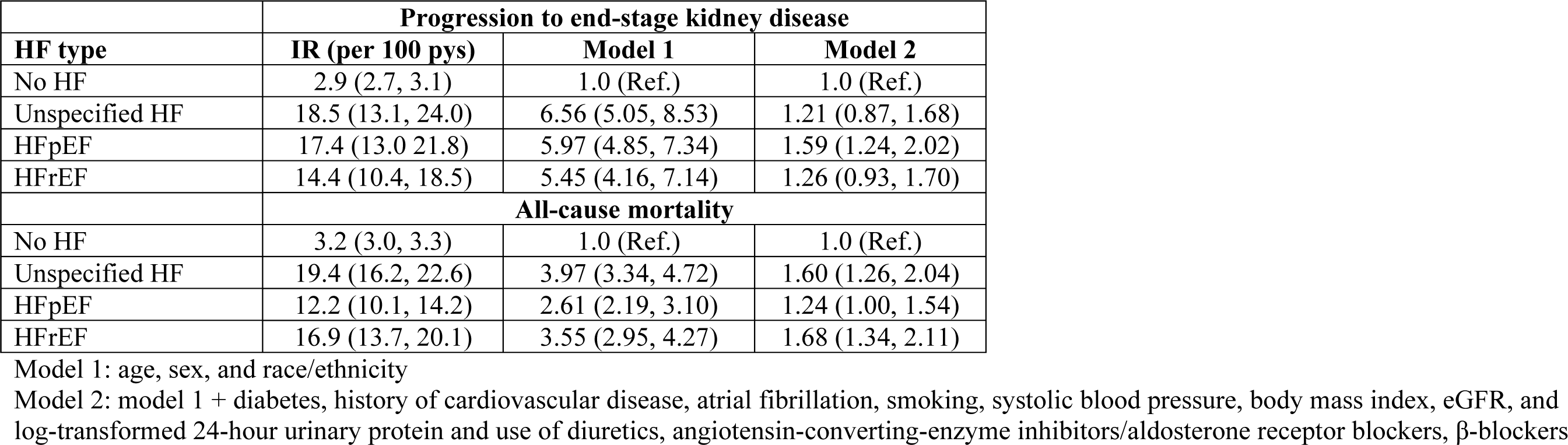
Association of heart failure subtypes with risk of end-stage kidney disease and mortality among participants with chronic kidney disease.

Among those who developed HFpEF, there was a strong graded association between lower eGFR category and higher risk of progression to ESKD (p-value for trend <0.0001); those with eGFR <30 ml/min/1.73m^2^ had greater that 5-fold higher risk of ESKD compared with those with eGFR>45 ml/min/1.73m^2^ after adjustment for possible confounders (**Figure 3**, **Supplemental Table 2 and Supplemental Figure 1a**). Similarly, among those who developed incident HFrEF, there was also a strong graded association between lower eGFR and risk of progression to ESKD (p-value for trend <0.0001, **Figure 3**, **Supplemental Table 3 and Supplemental Figure 2a**).

**Figure 3.**
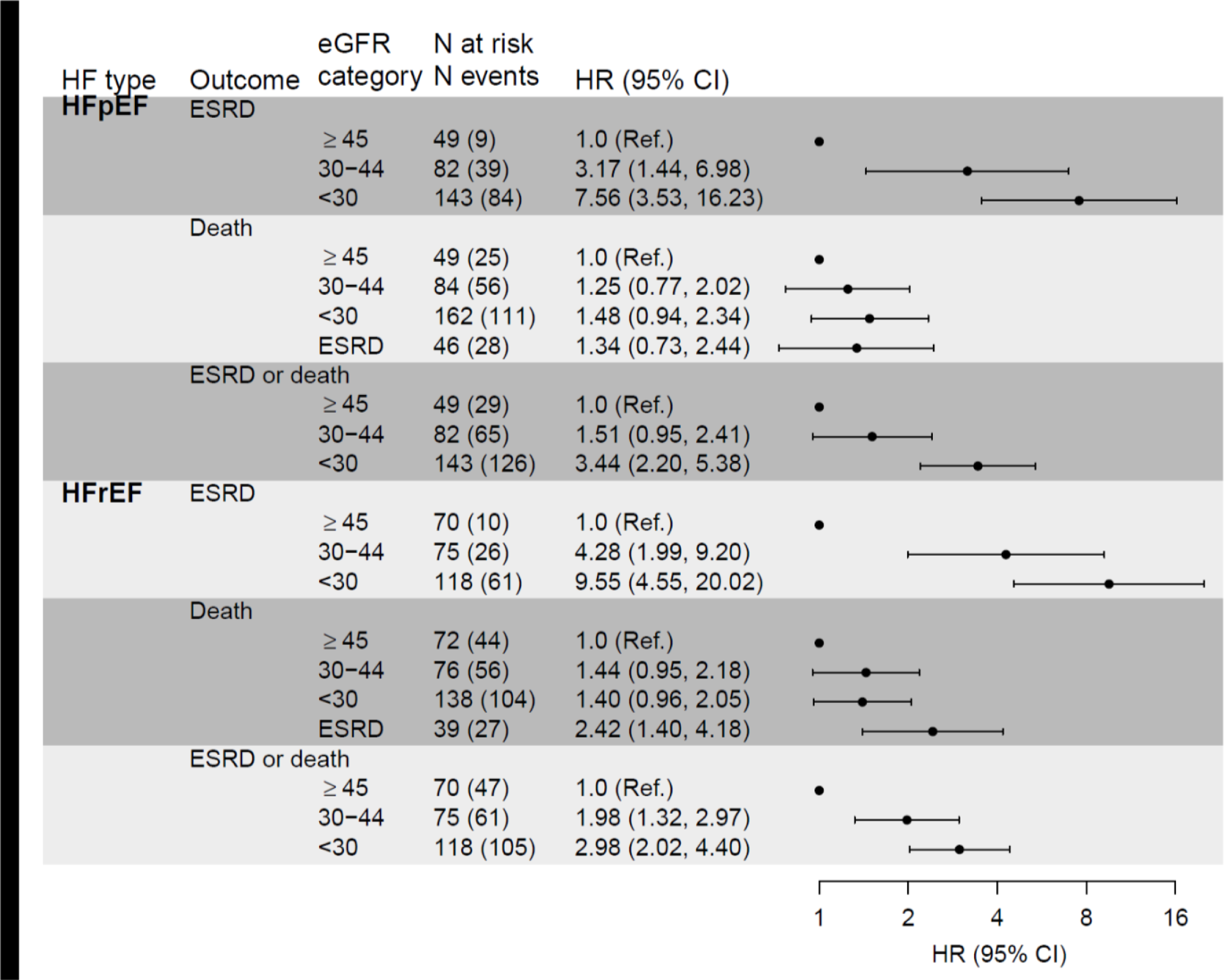
Association of heart failure with preserved (HFpEF) and reduced (HFrEF) ejection fraction with risk of progression to end-stage kidney disease and mortality across categories of estimated glomerular filtration rate (eGFR, ml/min/1.73m^2^)

### Association of HFpEF and HrpEF with mortality

Incidence rates of mortality were significantly higher in those who developed HF compared to those who did not develop HF. The median times to death among those with HFpEF and HFrEF were 3.2 (IQR 1.1-5.4) and 2.6 (IQR 1.1-5.4) years, respectively. In unadjusted models, incident HFpEF and HFrEF were associated with 3-4 fold higher risk of mortality. In models adjusted for demographics, comorbidity, measures of kidney function and medication use, both incident HFpEF and HFrEF retained significant, but largely attenuated, associations with mortality (HR 1.24 [95% CI: 1.00, 1.54] and HR 1.68 [95% CI: 1.34, 2.11], respectively) (**Table 2**). The risk of mortality was statistically greater with HFrEF vs. HFpEF (p-value = 0.02).

Among participants who developed incident HFpEF, 64% (220 of 344) died during follow-up, compared with 71% (231 of 325) of those with incident HFrEF. Risk of mortality did not appear to vary across eGFR categories among participants who developed incident HFpEF (p-value for trend = 0.54) (**Figure 3, Supplemental Table 2** and **Supplemental Figure 1b**), but did vary significantly among participants who developed HFrEF (p-value for trend = 0.01) (**Figure 3**, **Supplemental Table 3** and **Supplemental Figure 2b).**

## DISCUSSION

In this large, prospective cohort of adults with CKD, the rate of HFpEF hospitalizations was greater than that for HFrEF overall and across levels of eGFR. Additionally, there were many shared risk factors for both HF subtypes; however, higher inflammatory markers and alterations in mineral metabolism were more strongly associated with HFpEF. Statistically significant interactions by HF subtype were observed for male sex, blood pressure, heart rate and waist circumference. HFpEF and HFrEF were both associated with increased risk of progression to ESKD, with similar strengths of associations across both HF subtypes. Incident HFpEF and HFrEF were both characterized by extremely high mortality rates; however, the magnitude of association was greater for HFrEF than HFpEF compared with participants without HF. While much of HF management in CKD has focused on HFrEF, these data also highlight the large burden and poor outcomes associated with HFpEF in CKD.

In our study, participants with CKD experienced greater rates of HFpEF versus HFrEF hospitalizations (7 versus 5 per 1000 person years), consistent with studies from the general population.^21^ Prior studies had estimated that incidence rates of HFpEF to be ∼27 cases per 10,000 years;^22^ the rates of HFpEF in persons with CKD seen in our study were >10-fold higher. Other study populations of persons with CKD have also demonstrated higher rates of HFpEF versus HFrEF. Among patients with CKD and HF cared for at Kaiser Permanente of Southern California, 59% had HFpEF, 23% had HFrEF, and the remainder did not have EF available.^23^ Among HF patients in a Swedish Heart Failure Registry, CKD prevalence was 56% among those with HFpEF compared with 45% in those with HFrEF.^24^ Collectively, these data emphasize the need for increased attention to HFpEF in patients with CKD.

While there are many shared risk factors for HFpEF and HFrEF, there are also unique pathological features that make these two HF subtypes distinct.^25-27^ In our study older age, higher urine ACR, and history of atrial fibrillation and myocardial infarction were associated with risk of both HFpEF and HFrEF. Male sex, higher heart rate, history of COPD, smoking, lower eGFR, higher BMI lower waist circumference were more strongly associated with HFrEF, which are largely consistent with data from other non-CKD HF populations. In our study, statistically significant interactions by HF subtype were observed for male sex, blood pressure, heart rate and waist circumference. Studies of the general HF population also suggest that the mechanistic contributors for HFpEF include: myocardial fibrosis and cardiomyocyte hypertrophy induced from metabolic abnormalities, and endothelial dysfunction leading to microvascular disease and inflammation.^9^ Consistent with findings observed in the general HFpEF population, in our study of CKD patients, we observed significant associations of inflammatory markers (IL-6 and GDF-15) with risk of HFpEF. Unique to CKD, there were also significant associations of higher FGF-23 and phosphorus with risk of HFpEF (and not with HFrEF). Alterations in mineral metabolism have been linked with vascular calcification^28-30^ as well as myocyte hypertrophy,^31-33^ which likely explain the observed findings. Further investigation of CKD specific risk factors for HFpEF and HFrEF may lead the way for more tailored therapies.

We found that both HFpEF and HFrEF were significantly associated with increased risk for ESKD. In our previous study, we reported that frequency of HF hospitalizations was independently associated with greater risk of CKD progression and ESKD; of participants hospitalized for HF, 52% experienced significant CKD progression.^8^ Similarly, among patients with CKD in Canada, rates of ESKD were reported to be 4-to-14-fold higher among patients with previous HF hospitalizations.^34^ There are several possibilities to explain these findings. HF leads to hemodynamic changes, endothelial injury, inflammation and other processes which may further injure the kidneys.^35-37^ However, despite the high prevalence of HFpEF in patients with CKD, there are limited data on the risk of ESKD across HF subtypes; our study adds to the prognostic understanding of HFpEF in CKD. This is particularly important with the advent of exciting new therapies in the CKD-HF space. Trials have demonstrated robust and consistent benefit of sodium glucose co-transporter-2 inhibitors (SGLT2i) in decreasing rates of adverse kidney and HF outcomes for both HFpEF and HFrEF.^38-42^ SGLT2i should be part of the standard of care for patients with CKD and all HF subtypes for kidney protection.

In our study, 64% of patients who developed HF died. Both HFpEF and HFrEF were associated with higher risk of mortality; however, the risk was greater among patients with HFrEF. Previous studies have suggested similar associations. In a Swedish Heart Failure registry, CKD was more strongly associated with death in patients with HFrEF compared to HFpEF.^24^ Among patients with CKD in Kaiser of Southern California, the risk of mortality among those with HFrEF was also substantially higher than among those with HFpEF.^23^ In contrast, in a study of patients from multiple healthcare systems, the association of eGFR with risk of mortality was similar among patients with known HFpEF or HFrEF.^43^ Our study expands on these previous studies by studying a large, well-characterized national U.S. population with CKD. Given the extremely high proportion of post-HF hospitalization deaths in patients with CKD with both HFpEF and HFrEF, interventions to improve post-HF care should be prioritized in this high risk population.

These findings have important implications in the management of patients with CKD. Primary and secondary treatment of HF in CKD should be a public health priority, particularly with the exciting therapeutic advanced in recent years with medications such as SGLT2i and non-steroidal mineralocorticoid antagonists (MRAs). However, patients with CKD continue to be under-treated for HF, despite having higher rates of HF and worse HF-associated outcomes. A recent analysis found that patients with reduced eGFR at hospital discharge were significantly less likely to receive “triple therapy” with angiotensin-converting enzyme inhibitor/angiotensin receptor blocker/angiotensin receptor-neprilysin inhibitor + beta-blocker + MRAs, even at eGFR levels where such therapies are not contraindicated.^44^ In part, this may be due to concern about adverse effects. For example, many therapies (e.g. SGLT2i and RAASi) may lead to a ‘eGFR dip;’ however studies have shown that despite this dip, there are clear benefits to these medications in persons with CKD.^45,46^ With new therapies available, there should be increased focus on new strategies for implementation of HF therapies in persons with CKD.

Our study had several strengths. We studied a large, multi-center, well-characterized, U.S. based CKD population specifically designed to study cardiovascular complications with extensive longitudinal follow-up. All HF hospitalizations that occurred during CRIC follow-up were adjudicated using standardized criteria. We recognize a few limitations as well. We were not able to determine whether acute kidney injury occurred during the HF hospitalizations, which may have contributed to subsequent CKD progression. Ejection fraction data to determine HF subtype were not available in all participants. We did not have detailed data on whether certain medications (e.g. RAAS inhibitors or diuretics) were held or doses adjusted after the hospitalizations since medication use was only ascertained at six months intervals per the CRIC study protocol. Data on SGLT2i use were not available since this cohort largely (?) predated availability of these medications. While the CRIC adjudication process is known to identify >90% of hospitalizations, some HF hospitalizations may have been missed. This was a clinic-based population of research volunteers, so it may not be generalizable to all CKD patients. In this observational study, we cannot determine causality, and we cannot exclude reverse causality.

In conclusion, in a large U.S. CKD population, the rates of HFpEF hospitalizations were approximately 30% greater than that of HFrEF. There were many shared risk factors for both HF subtypes, however some unique risk factors were identified for HFpEF and HFrEF in this population of CKD patients. Both types of HF had similar associations with risk of ESKD in this population; with substantial risk amongst those with lower eGFR. Almost 2/3 of persons who developed HF died during study follow-up; however, there was a stronger association of HFrEF with mortality compared with HFpEF. Prevention and treatment of HFpEF as well as HFrEF should be a central priority to improve clinical outcomes in patients with CKD.

### Disclaimer

This article was not prepared in collaboration with investigators of the Chronic Renal Insufficiency Cohort (CRIC) study and does not necessarily reflect the opinions or views of the CRIC study investigators, the National Institute of Diabetes and Digestive and Kidney Diseases (NIDDK) central repositories, or the NIDDK.

### Additional Information

The CRIC was conducted by the CRIC study investigators and supported by the NIDDK. The data from the CRIC reported here were supplied by the NIDDK central repositories.

## Data Availability

all data available through NIDDK repository

---

**Supplemental Table 1.** Demographic characteristics of the study population (N = 3557)

|  | **Overall** |
| --- | --- |
| **N** | 3557 |
| **Demographics** |  |
| Age | 57.4 (11.2) |
| Male | 1941 (55) |
| Race/ethnicity |  |
| Non-Hispanic white | 1521 (43) |
| Non-Hispanic black | 1433 (40) |
| Hispanic | 460 (13) |
| Other | 143 (4) |
| Income |  |
| <\$20K | 1057 (30) |
| \$20-<\$50K | 873 (25) |
| \$50-<\$100K | 687 (19) |
| ≥\$100K | 377 (11) |
| Education |  |
| ≤6<sup>th</sup> grade | 194 (5) |
| 7<sup>th</sup>-12<sup>th</sup> grade | 538 (15) |
| HS graduate | 648 (18) |
| Technical school | 174 (5) |
| Some college | 846 (24) |
| College graduate | 671 (19) |
| Graduate or professional school | 485 (14) |
| Alcohol use | 2278 (64) |
| **Physical characteristics** |  |
| BMI (kg/m<sup>2</sup>) | 31.9 (7.7) |
| Waist circumference (cm) | 105.3 (17.4) |
| SBP (mmHg) | 128.5 (22.0) |
| DBP (mmHg) | 71.9 (12.6) |
| **Comorbidity** |  |
| Atrial fibrillation | 482 (14) |
| COPD | 98 (3) |
| Diabetes | 1645 (46) |
| MI | 626 (18) |
| PVD | 202 (6) |
| Stroke | 320 (9) |
| Smoking | 463 (13) |
| Total METs | 167 (111-252) |
| **Laboratory** |  |
| APOA1 | 137.4 (29.9) |
| Triglycerides | 155.5 (115.2) |
| Total cholesterol | 185.0 (45.5) |
| HbA1c | 6.6 (1.5) |
| Hemoglobin | 12.6 (1.8) |
| Glucose | 114.1 (50.8) |
| Serum albumin | 3.9 (0.5) |
| Sodium | 139.2 (3.1) |
| Serum creatinine | 1.7 (0.6) |
| Serum cystatin-C | 1.5 (0.5) |
| eGFR (CKD-EPI) | 50.2 (19.8) |
| Urine volume from 24-hour test | 2079 (845) |
| Total 24-hour urine protein, g/day | 0.2 (0.1-0.9) |
| 24-hour urine sodium | 161.2 (77.1) |
| 24-hour urine potassium | 55.1 (26.3) |
| hsCRP | 2.5 (1.0-6.3) |
| IL-10 > 0, N (%) | 539 (15) |
| IL-6 | 1.8 (1.1-3.0) |
| **Mineral metabolism** |  |
| Calcium | 9.2 (0.5) |
| Phosphorus | 3.7 (0.7) |
| PTH | 52.0 (34.0-84.8) |
| FGF-23 | 138.7 (93.5-224.2) |
| **Electrocardiogram** |  |
| Heart rate | 65.2 (11.8) |
| QRS interval | 92 (86-100) |
| QT interval | 404 (382-430) |
| PR interval | 168 (152-186) |
| **Symptoms** |  |
| Year 1 KCCQ score | 91 (73-99) |
| KDQOL symptoms score | 89 (77-95) |
| KDQOL burden score | 94 (69-100) |
| KDQOL effects score | 97 (88-100) |
| **Depression** |  |
| Mild | 2957 (83) |
| Moderate | 465 (13) |
| Severe | 91 (3) |
| **Cardiac biomarkers** |  |
| NT-proBNP | 123 (45-327) |
| hsTnT | 13.9 (8.3-24.4) |
| Galectin-3 | 14.0 (10.0-19.2) |
| GDF-15 | 1418 (967-2124) |
| sST2 | 15.3 (11.3-20.8) |

**Supplemental Table 2.** Association of eGFR with risk of end-stage kidney disease and mortality among participants with incident HFpEF.

|  | **Progression to end-stage kidney disease** |  |  |  |  |
| --- | --- | --- | --- | --- | --- |
| eGFR | **N at risk**<br>**(N events)** | **IR (per 100 pys)** | **Unadjusted** | **Model 1** | **Model 2** |
| Overall | 274 (132) | 17.5 (14.3, 21.7) |  |  |  |
| eGFR $\geq 45$ | 49 (9) | 3.4 (1.5, 6.1) | 1.0 (Ref.) | 1.0 (Ref.) | 1.0 (Ref.) |
| eGFR 30-44 | 82 (39) | 14.0 (9.8, 20.0) | 2.24 (1.20, 4.20) | 2.82 (1.47, 5.40) | 3.10 (1.41, 6.84) |
| eGFR $< 30$ | 143 (84) | 38.6 (29.9, 50.7) | 5.09 (2.78, 9.29) | 7.19 (3.84, 13.44) | 7.55 (3.51, 16.28) |
|  | **All-cause mortality** |  |  |  |  |
| Overall | 341 (220) | 13.1 (11.4, 15.0) |  |  |  |
| eGFR $\geq 45$ | 49 (25) | 8.6 (5.4, 13.1) | 1.0 (Ref.) | 1.0 (Ref.) | 1.0 (Ref.) |
| eGFR 30-44 | 84 (56) | 12.5 (9.7, 16.3) | 1.83 (1.14, 2.93) | 1.65 (1.02, 2.66) | 1.24 (0.76, 2.01) |
| eGFR $< 30$ | 162 (111) | 14.6 (12.1, 17.8) | 1.80 (1.14, 2.84) | 1.61 (1.02, 2.56) | 1.47 (0.93, 2.34) |
| ESRD | 46 (28) | 16.5 (10.8, 24.3) | 2.08 (1.19, 3.62) | 1.79 (1.01, 3.16) | 1.32 (0.71, 2.45) |
|  | **Composite of progression to end-stage kidney disease and all-cause mortality** |  |  |  |  |
| Overall | 274 (220) | 29.4 (24.6, 34.8) |  |  |  |
| eGFR $\geq 45$ | 49 (29) | 11.4 (7.5, 16.6) | 1.0 (Ref.) | 1.0 (Ref.) | 1.0 (Ref.) |
| eGFR 30-44 | 82 (65) | 23.4 (17.7, 31.5) | 2.11 (1.37, 3.26) | 2.24 (1.43, 3.51) | 1.56 (0.97, 2.49) |
| eGFR $< 30$ | 143 (126) | 58.4 (47.1, 73.4) | 3.71 (2.41, 5.69) | 4.21 (2.70, 6.57) | 3.50 (2.21, 5.52) |

Model 1: age, sex, and race/ethnicity
Model 2: model 1 + diabetes, history of cardiovascular disease, atrial fibrillation, smoking, systolic blood pressure, body mass index, and log-transformed 24-hour urine protein and use of diuretics, angiotensin-converting-enzyme inhibitors/aldosterone receptor blockers, β-blockers

**Supplemental Table 3.** Association of eGFR with risk of end-stage kidney disease and mortality among participants with incident HFrEF.

|  | **Progression to end-stage kidney disease** |  |  |  |  |
| --- | --- | --- | --- | --- | --- |
| eGFR | **N at risk**<br>**(N events)** | **IR (per 100 pys)** | **Unadjusted** | **Model 1** | **Model 2** |
| Overall | 263 (97) | 13.2 (10.6, 16.3) |  |  |  |
| eGFR≥45 | 70 (10) | 3.1 (1.4, 5.0) | 1.0 (Ref.) | 1.0 (Ref.) | 1.0 (Ref.) |
| eGFR 30-44 | 75 (26) | 14.3 (9.2, 20.4) | 3.28 (1.60, 6.72) | 3.45 (1.67, 7.11) | 3.53 (1.61, 7.73) |
| eGFR <30 | 118 (61) | 26.2 (18.7, 36.5) | 7.37 (3.73, 14.55) | 8.55 (4.31, 16.97) | 9.30 (4.41, 19.6) |
|  | **All-cause mortality** |  |  |  |  |
| Overall | 325 (231) | 17.8 (15.6, 20.5) |  |  |  |
| eGFR≥45 | 72 (44) | 12.5 (9.4, 16.8) | 1.0 (Ref.) | 1.0 (Ref.) | 1.0 (Ref.) |
| eGFR 30-44 | 76 (56) | 21.4 (16.5, 28.1) | 1.68 (1.13, 2.48) | 1.62 (1.09, 2.40) | 1.46 (0.95, 2.24) |
| eGFR <30 | 138 (104) | 17.9 (14.7, 22.0) | 1.49 (1.03, 2.16) | 1.56 (1.08, 2.26) | 1.39 (0.94, 2.04) |
| ESRD | 39 (27) | 26.7 (16.5, 45.6) | 2.17 (1.33, 3.55) | 2.32 (1.41, 3.80) | 2.39 (1.35, 4.23) |
|  | **Composite of progression to end-stage kidney disease and all-cause mortality** |  |  |  |  |
| Overall | 263 (213) | 29.2 (24.9, 34.1) |  |  |  |
| eGFR ≥45 | 70 (47) | 14.9 (11.2, 19.3) | 1.0 (Ref.) | 1.0 (Ref.) | 1.0 (Ref.) |
| eGFR 30-44 | 75 (61) | 33.7 (25.8, 44.4) | 1.79 (1.22, 2.62) | 1.69 (1.15, 2.48) | 1.90 (1.26, 2.88) |
| eGFR <30 | 118 (105) | 45.5 (34.6, 59.5) | 3.08 (2.14, 4.42) | 3.26 (2.27, 4.70) | 2.90 (1.95, 4.30) |

Model 1: age, sex, and race/ethnicity
Model 2: model 1 + diabetes, history of cardiovascular disease, atrial fibrillation, smoking, systolic blood pressure, body mass index, and log-transformed 24-hour urine protein and use of diuretics, angiotensin-converting-enzyme inhibitors/aldosterone receptor blockers, β-blockers

**Supplemental Figure 1.**
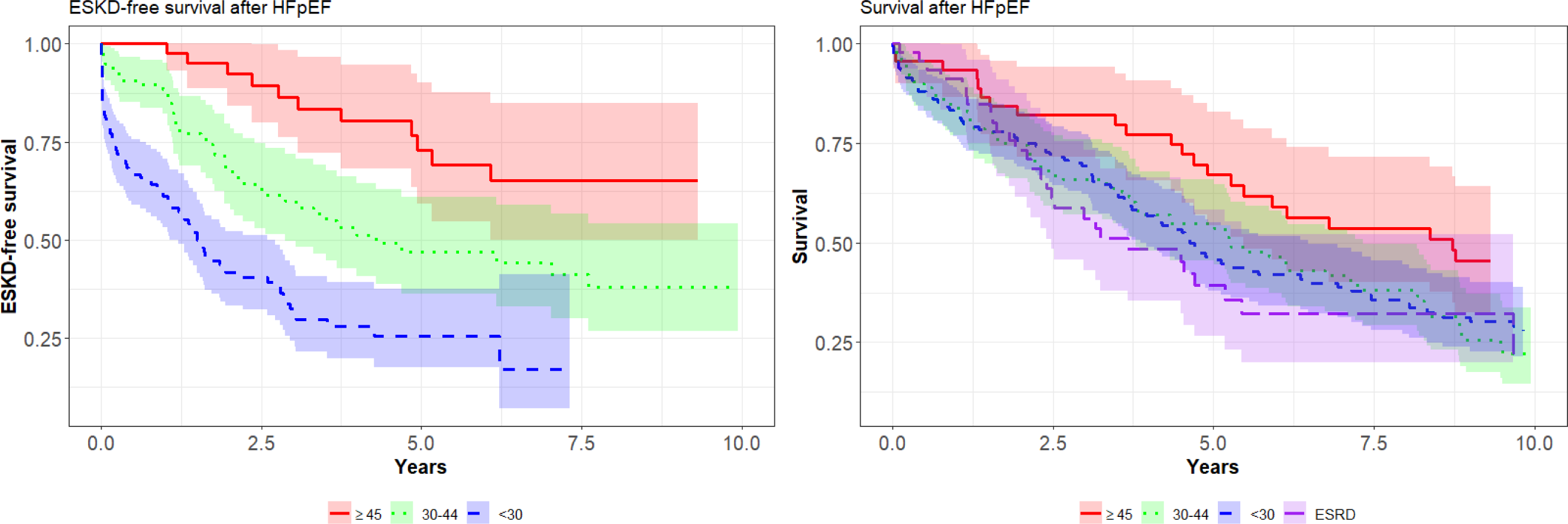
(a) End-stage kidney disease (ESKD) free survival and (b) overall survival after incident heart failure with preserved ejection fraction (HFpEF)

**Supplemental Figure 2.**
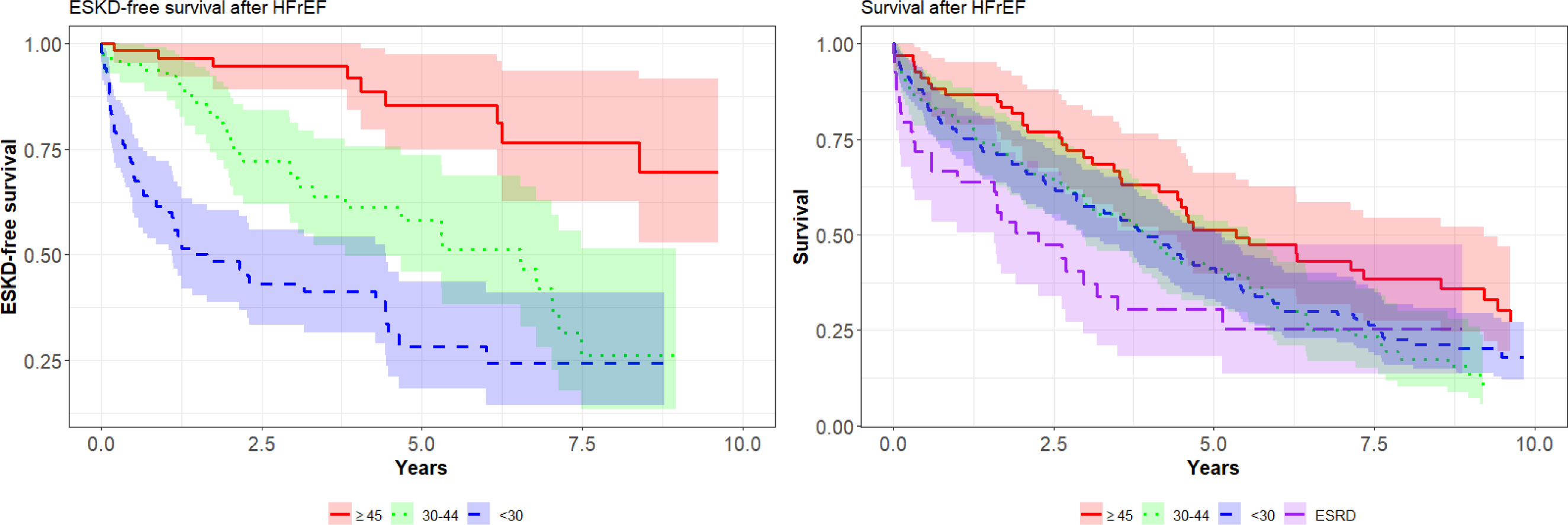
(a) End-stage kidney disease (ESKD) free survival and (b) overall survival after incident heart failure with reduced ejection fraction (HFrEF)

## REFERENCES

1. Bansal N, Katz R, Robinson-Cohen C, Odden MC, Dalrymple L, Shlipak MG, Sarnak MJ, Siscovick DS, Zelnick L, Psaty BM, et al. Absolute Rates of Heart Failure, Coronary Heart Disease, and Stroke in Chronic Kidney Disease: An Analysis of 3 Community-Based Cohort Studies. JAMA Cardiol. 2017;2:314–318. doi: 10.1001/jamacardio.2016.4652

2. Kottgen A, Russell SD, Loehr LR, Crainiceanu CM, Rosamond WD, Chang PP, Chambless LE, Coresh J. Reduced kidney function as a risk factor for incident heart failure: the atherosclerosis risk in communities (ARIC) study. J Am Soc Nephrol. 2007;18:1307–1315. doi: 10.1681/asn.2006101159

3. Go AS, Chertow GM, Fan D, McCulloch CE, Hsu CY. Chronic kidney disease and the risks of death, cardiovascular events, and hospitalization. N Engl J Med. 2004;351:1296–1305. doi: 10.1056/NEJMoa041031 [doi] 351/13/1296 [pii]

4. Bansal N, Katz R, Robinson-Cohen C, Odden MC, Dalrymple L, Shlipak MG, Sarnak MJ, Siscovick DS, Zelnick L, Psaty BM, et al. Absolute Rates of Heart Failure, Coronary Heart Disease, and Stroke in Chronic Kidney Disease: An Analysis of 3 Community-Based Cohort Studies. JAMA Cardiol. 2016. doi: 10.1001/jamacardio.2016.4652

5. 2016 USRDS Annual Data Report: Epidemiology of kidney disease in the United States. In; 2016.

6. Harel Z, Wald R, McArthur E, Chertow GM, Harel S, Gruneir A, Fischer HD, Garg AX, Perl J, Nash DM, et al. Rehospitalizations and Emergency Department Visits after Hospital Discharge in Patients Receiving Maintenance Hemodialysis. J Am Soc Nephrol. 2015;26:3141–3150. doi: 10.1681/asn.2014060614

7. Schefold JC, Filippatos G, Hasenfuss G, Anker SD, von Haehling S. Heart failure and kidney dysfunction: epidemiology, mechanisms and management. Nature Reviews Nephrology. 2016;12:610–623. doi: 10.1038/nrneph.2016.113

8. Bansal N, Zelnick L, Bhat Z, Dobre M, He J, Lash J, Jaar B, Mehta R, Raj D, Rincon-Choles H, et al. Burden and Outcomes of Heart Failure Hospitalizations in Adults With Chronic Kidney Disease. J Am Coll Cardiol. 2019;73:2691–2700. doi: 10.1016/j.jacc.2019.02.071

9. Borlaug BA, Sharma K, Shah SJ, Ho JE. Heart Failure With Preserved Ejection Fraction: JACC Scientific Statement. J Am Coll Cardiol. 2023;81:1810–1834. doi: 10.1016/j.jacc.2023.01.049

10. Desai AS, Lam CSP, McMurray JJV, Redfield MM. How to Manage Heart Failure With Preserved Ejection Fraction: Practical Guidance for Clinicians. JACC Heart Fail. 2023;11:619–636. doi: 10.1016/j.jchf.2023.03.011

11. Feldman HI, Appel LJ, Chertow GM, Cifelli D, Cizman B, Daugirdas J, Fink JC, Franklin-Becker ED, Go AS, Hamm LL, et al. The Chronic Renal Insufficiency Cohort (CRIC) Study: Design and Methods. J Am Soc Nephrol. 2003;14:S148–153.

12. Lash JP, Go AS, Appel LJ, He J, Ojo A, Rahman M, Townsend RR, Xie D, Cifelli D, Cohan J, et al. Chronic Renal Insufficiency Cohort (CRIC) Study: Baseline Characteristics and Associations with Kidney Function. Clin J Am Soc Nephrol. 2009;4:1302–1311. doi: CJN.00070109 [pii] 10.2215/CJN.00070109 [doi]

13. Bansal N, Roy J, Chen HY, Deo R, Dobre M, Fischer MJ, Foster E, Go AS, He J, Keane MG, et al. Evolution of Echocardiographic Measures of Cardiac Disease From CKD to ESRD and Risk of All-Cause Mortality: Findings From the CRIC Study. Am J Kidney Dis. 2018. doi: 10.1053/j.ajkd.2018.02.363

14. Bansal N, Keane M, Delafontaine P, Dries D, Foster E, Gadegbeku CA, Go AS, Hamm LL, Kusek JW, Ojo AO, et al. A longitudinal study of left ventricular function and structure from CKD to ESRD: the CRIC study. Clin J Am Soc Nephrol. 2013;8:355–362. doi: 10.2215/cjn.06020612

15. National Center for Health Statistics (NCHS). National Health and Nutrition Examination Survey Anthropometry Procedures Manual. Centers for Disease Control and Prevention [serial online] 2000.

16. Joffe M, Hsu CY, Feldman HI, Weir M, Landis JR, Hamm LL, Chronic Renal Insufficiency Cohort Study G. Variability of creatinine measurements in clinical laboratories: results from the CRIC study. Am J Nephrol. 2010;31:426–434. doi: 10.1159/000296250

17. Levey AS, Coresh J, Greene T, Marsh J, Stevens LA, Kusek JW, Van Lente F, Chronic Kidney Disease Epidemiology C. Expressing the Modification of Diet in Renal Disease Study equation for estimating glomerular filtration rate with standardized serum creatinine values. Clin Chem. 2007;53:766–772. doi: 10.1373/clinchem.2006.077180

18. Inker LA, Eneanya ND, Coresh J, Tighiouart H, Wang D, Sang Y, Crews DC, Doria A, Estrella MM, Froissart M, et al. New Creatinine- and Cystatin C-Based Equations to Estimate GFR without Race. N Engl J Med. 2021;385:1737–1749. doi: 10.1056/NEJMoa2102953

19. http://www.kdigo.org/clinical_practice_guidelines/CKD.php.

20. Wei LJ, Lin DY, Weissfeld L. Regression Analysis of Multivariate Incomplete Failure Time Data by Modeling Marginal Distributions. Journal of the American Statistical Association. 1989;84:1065–1073. doi: 10.2307/2290084

21. Gerber Y, Weston SA, Redfield MM, Chamberlain AM, Manemann SM, Jiang R, Killian JM, Roger VL. A contemporary appraisal of the heart failure epidemic in Olmsted County, Minnesota, 2000 to 2010. JAMA Intern Med. 2015;175:996–1004. doi: 10.1001/jamainternmed.2015.0924

22. Bhambhani V, Kizer JR, Lima JAC, van der Harst P, Bahrami H, Nayor M, de Filippi CR, Enserro D, Blaha MJ, Cushman M, et al. Predictors and outcomes of heart failure with mid-range ejection fraction. Eur J Heart Fail. 2018;20:651–659. doi: 10.1002/ejhf.1091

23. Yu AS, Pak KJ, Zhou H, Shaw SF, Shi J, Broder BI, Sim JJ. All-Cause and Cardiovascular-Related Mortality in CKD Patients With and Without Heart Failure: A Population-Based Cohort Study in Kaiser Permanente Southern California. Kidney Med. 2023;5:100624. doi: 10.1016/j.xkme.2023.100624

24. Löfman I, Szummer K, Dahlström U, Jernberg T, Lund LH. Associations with and prognostic impact of chronic kidney disease in heart failure with preserved, mid-range, and reduced ejection fraction. Eur J Heart Fail. 2017;19:1606–1614. doi: 10.1002/ejhf.821

25. Peters AE, Tromp J, Shah SJ, Lam CSP, Lewis GD, Borlaug BA, Sharma K, Pandey A, Sweitzer NK, Kitzman DW, et al. Phenomapping in heart failure with preserved ejection fraction: insights, limitations, and future directions. Cardiovasc Res. 2023;118:3403–3415. doi: 10.1093/cvr/cvac179

26. Shah SJ, Katz DH, Deo RC. Phenotypic spectrum of heart failure with preserved ejection fraction. Heart Fail Clin. 2014;10:407–418. doi: 10.1016/j.hfc.2014.04.008

27. Shah SJ, Katz DH, Selvaraj S, Burke MA, Yancy CW, Gheorghiade M, Bonow RO, Huang CC, Deo RC. Phenomapping for novel classification of heart failure with preserved ejection fraction. Circulation. 2015;131:269–279. doi: 10.1161/circulationaha.114.010637

28. Doshi SM, Wish JB. Past, Present, and Future of Phosphate Management. Kidney Int Rep. 2022;7:688–698. doi: 10.1016/j.ekir.2022.01.1055

29. Ray M, Jovanovich A. Mineral Bone Abnormalities and Vascular Calcifications. Adv Chronic Kidney Dis. 2019;26:409–416. doi: 10.1053/j.ackd.2019.09.004

30. Villa-Bellosta R. Vascular Calcification: Key Roles of Phosphate and Pyrophosphate. Int J Mol Sci. 2021;22. doi: 10.3390/ijms222413536

31. Grabner A, Faul C. The role of fibroblast growth factor 23 and Klotho in uremic cardiomyopathy. Curr Opin Nephrol Hypertens. 2016;25:314–324. doi: 10.1097/mnh.0000000000000231

32. Navarro-García JA, Fernández-Velasco M, Delgado C, Delgado JF, Kuro OM, Ruilope LM, Ruiz-Hurtado G. PTH, vitamin D, and the FGF-23-klotho axis and heart: Going beyond the confines of nephrology. Eur J Clin Invest. 2018;48. doi: 10.1111/eci.12902

33. Faul C, Amaral AP, Oskouei B, Hu MC, Sloan A, Isakova T, Gutiérrez OM, Aguillon-Prada R, Lincoln J, Hare JM, et al. FGF23 induces left ventricular hypertrophy. J Clin Invest. 2011;121:4393–4408. doi: 10.1172/jci46122

34. Sud M, Tangri N, Pintilie M, Levey AS, Naimark DM. ESRD and death after heart failure in CKD. J Am Soc Nephrol. 2015;26:715–722. doi: 10.1681/asn.2014030253

35. He J, Shlipak M, Anderson A, Roy JA, Feldman HI, Kallem RR, Kanthety R, Kusek JW, Ojo A, Rahman M, et al. Risk Factors for Heart Failure in Patients With Chronic Kidney Disease: The CRIC (Chronic Renal Insufficiency Cohort) Study. J Am Heart Assoc. 2017;6. doi: 10.1161/jaha.116.005336

36. Michowitz Y, Goldstein E, Wexler D, Sheps D, Keren G, George J. Circulating endothelial progenitor cells and clinical outcome in patients with congestive heart failure. Heart. 2007;93:1046–1050. doi: 10.1136/hrt.2006.102657

37. Bansal N, Katz R, Dalrymple L, de Boer I, DeFilippi C, Kestenbaum B, Park M, Sarnak M, Seliger S, Shlipak M. NT-proBNP and troponin T and risk of rapid kidney function decline and incident CKD in elderly adults. Clin J Am Soc Nephrol. 2015;10:205–214. doi: 10.2215/cjn.04910514

38. Bhatt DL, Szarek M, Steg PG, Cannon CP, Leiter LA, McGuire DK, Lewis JB, Riddle MC, Voors AA, Metra M, et al. Sotagliflozin in Patients with Diabetes and Recent Worsening Heart Failure. N Engl J Med. 2021;384:117–128. doi: 10.1056/NEJMoa2030183

39. McMurray JJV, Solomon SD, Inzucchi SE, Køber L, Kosiborod MN, Martinez FA, Ponikowski P, Sabatine MS, Anand IS, Bělohlávek J, et al. Dapagliflozin in Patients with Heart Failure and Reduced Ejection Fraction. N Engl J Med. 2019;381:1995–2008. doi: 10.1056/NEJMoa1911303

40. Packer M, Anker SD, Butler J, Filippatos G, Pocock SJ, Carson P, Januzzi J, Verma S, Tsutsui H, Brueckmann M, et al. Cardiovascular and Renal Outcomes with Empagliflozin in Heart Failure. N Engl J Med. 2020;383:1413–1424. doi: 10.1056/NEJMoa2022190

41. Perkovic V, Jardine MJ, Neal B, Bompoint S, Heerspink HJL, Charytan DM, Edwards R, Agarwal R, Bakris G, Bull S, et al. Canagliflozin and Renal Outcomes in Type 2 Diabetes and Nephropathy. N Engl J Med. 2019;380:2295–2306. doi: 10.1056/NEJMoa1811744

42. Solomon SD, McMurray JJV, Claggett B, de Boer RA, DeMets D, Hernandez AF, Inzucchi SE, Kosiborod MN, Lam CSP, Martinez F, et al. Dapagliflozin in Heart Failure with Mildly Reduced or Preserved Ejection Fraction. N Engl J Med. 2022;387:1089–1098. doi: 10.1056/NEJMoa2206286

43. Smith DH, Thorp ML, Gurwitz JH, McManus DD, Goldberg RJ, Allen LA, Hsu G, Sung SH, Magid DJ, Go AS. Chronic kidney disease and outcomes in heart failure with preserved versus reduced ejection fraction: the Cardiovascular Research Network PRESERVE Study. Circulation Cardiovascular quality and outcomes. 2013;6:333–342. doi: 10.1161/circoutcomes.113.000221

44. Patel RB, Fonarow GC, Greene SJ, Zhang S, Alhanti B, DeVore AD, Butler J, Heidenreich PA, Huang JC, Kittleson MM, et al. Kidney Function and Outcomes in Patients Hospitalized With Heart Failure. J Am Coll Cardiol. 2021;78:330–343. doi: 10.1016/j.jacc.2021.05.002

45. Xie Y, Bowe B, Gibson AK, McGill JB, Maddukuri G, Al-Aly Z. Clinical Implications of Estimated Glomerular Filtration Rate Dip Following Sodium-Glucose Cotransporter-2 Inhibitor Initiation on Cardiovascular and Kidney Outcomes. J Am Heart Assoc. 2021;10:e020237. doi: 10.1161/jaha.120.020237

46. McCallum W, Tighiouart H, Ku E, Salem D, Sarnak MJ. Trends in Kidney Function Outcomes Following RAAS Inhibition in Patients With Heart Failure With Reduced Ejection Fraction. Am J Kidney Dis. 2020;75:21–29. doi: 10.1053/j.ajkd.2019.05.010

